# TzanckNet: A convolutional neural network to identify cells in the cytology of erosive-vesiculobullous diseases

**DOI:** 10.1101/2020.06.22.20137570

**Authors:** Mehmet Alican Noyan, Murat Durdu, Ali Haydar Eskiocak

**Affiliations:** Ipsumio B.V., High Tech Campus, 5656 AE Eindhoven, The Netherlands; Department of Dermatology, Başkent University Medical School, Adana Dr. Turgut Noyan Application and Research Center, Adana, Turkey

**Keywords:** Tzanck Smear test, Deep Learning, Cutaneous Cytology, Vesiculobullous Skin Disease

## Abstract

Tzanck smear test is a low-cost, rapid and reliable tool which can be used for the diagnosis of many erosive-vesiculobullous, tumoral and granulomatous diseases. Currently its use is limited mainly due to lack of experience in interpretation of the smears. We developed a deep learning model, TzanckNet, that can identify cells in Tzanck smear test findings. TzanckNet was trained on a retrospective development dataset of 2260 Tzanck smear images collected between December 2006 – December 2019. The finalized model was evaluated using a prospective validation dataset of 359 Tzanck smear images collected from 15 patients during January 2020. It is designed to recognize six cell types (acantholytic cells, eosinophils, hypha, multinucleated giant cells, normal keratinocytes and tadpole cells). For 359 images and 6 cell types, TzanckNet made 2154 predictions. The accuracy was 94.3 % (95% CI 93.4 to 95.3), the sensitivity was 83.7 % (95% CI 80.3 to 87.0) and the specificity was 97.3 % (95% CI 96.5 to 98.1). The area under the receiver operating characteristic curve was 0.974. Our results show that TzanckNet has the potential to lower the experience barrier needed to use this test, broadening its user base, and hence improving patient well-being.

## Introduction

A dermatologist is expected to provide an accurate diagnosis. Accuracy, however, is hardly the only criterion for choosing the right diagnostic test. The cost has always been an important concern and is even more so today with rising healthcare costs in certain countries. Regulations add another dimension to consider. Many tests that are taken for granted in some clinics, might not be available in resource-constrained settings. Patient discomfort is yet another consideration. Therefore, it is hard to find a diagnostic test that checks all the boxes.

Tzanck smear test, a dermatocytological diagnostic tool first described in 1947^1^, costs less than a dollar, gives results in less than an hour, causes minimal patient discomfort, does not require anesthesia and is accurate.^2^ It requires simple tools that almost all the clinics in the world can access – a microscope slide, an immersion oil, a scalpel, cytological stains and a microscope. With these advantages, one would expect it to be indispensable for any dermatologist’s toolbox.

Surprisingly, the reality is quite the opposite. Although this dermatologist friendly method can be used for the diagnosis of many erosive-vesiculobullous, tumoral and granulomatous diseases, its use is usually limited to herpes virus infections in daily dermatology practice.^2^ In some clinics, this test is not used even for the diagnosis of herpes virus infections; instead patients receive local anesthesia followed by a skin biopsy.^3^ Furthermore, when folliculitis patients are treated without cytological examination, patients with viral, parasitic and fungal folliculitis may receive unnecessary antibiotic treatments for years.^4^ One of the reasons for this is the lack of experience in using this test. Efforts have been made in the past decade to reintroduce Tzanck smear test5– ^10^ which has resulted in revived interest.^11–13^ Still, this diagnostic tool is nowhere near its full potential. The main textbooks of dermatopathology do not include dedicated sections for cytology or Tzanck smear.^14,15^

Artificial intelligence has long been used to tackle challenges in healthcare^16–19^, with growing attention in the last years. It has found wide ranging roles from automating menial tasks^20^ to optimizing medical resources.^21^ The spotlight, however, is on deep learning models showing expert level performance on certain medical skills – even surpassing humans in some instances.^22– 25^ Analyzing medical images constitute an important part of these skills^26^ because the development of deep learning has been strongly tied to image processing.^27^ Artificial intelligence models have been used in dermatology^19,23,28–31^, but, to the best of our knowledge, they have never been applied to cutaneous cytology or evaluating Tzanck smear findings.

The aim of this study was to develop a deep neural network, called TzanckNet, for recognizing cells in Tzanck smear test findings of erosive-vesiculobullous diseases. It was designed to recognize six cell types related to diseases such as herpetic infections, pemphigus, and spongiotic dermatitis (Figure 1). The model was developed using a retrospective dataset and validated clinically using a prospective dataset.

**Figure 1.**
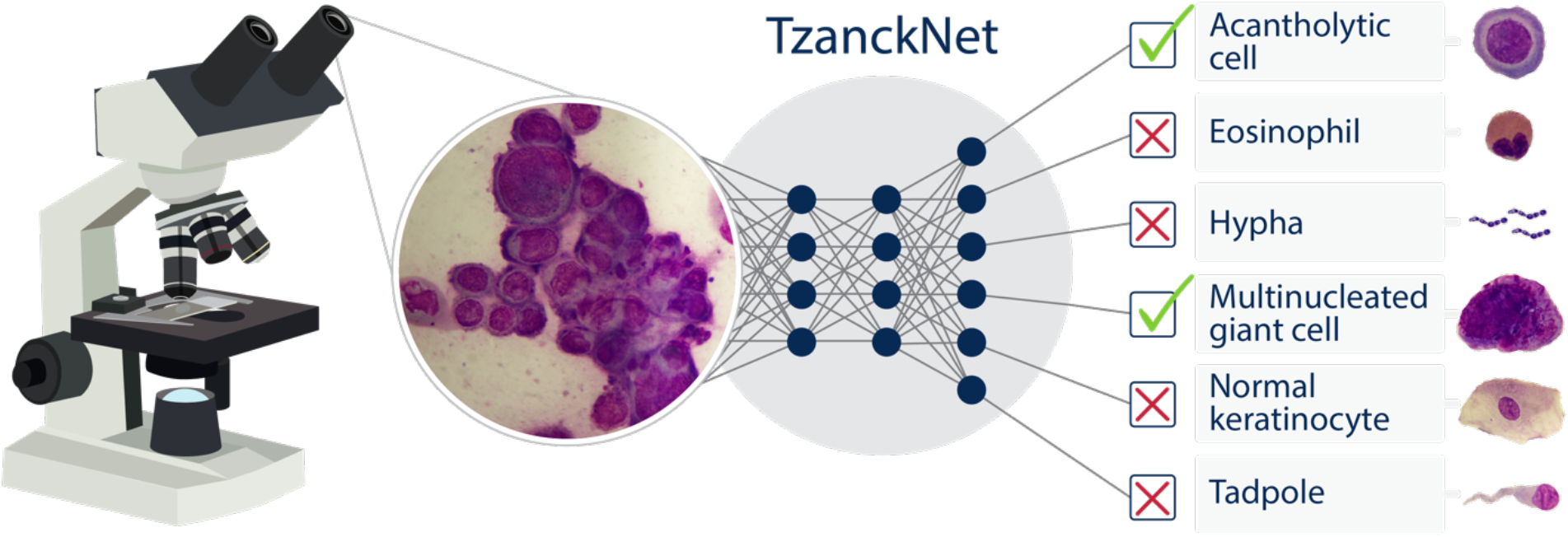
A schematic showing how the proposed TzanckNet works. TzanckNet accepts a Tzanck smear image as an input and outputs which cell types among six cell types are present and absent in the image. As an example, a Tzanck smear image from a patient with herpetic infection goes into the TzanckNet and the network predicts that there are acantholytic and multinucleated giant cells in the image, the remaining four cell types does not exist in the image. On the right-hand side of the figure, an example image from each cell type is presented.

## Methods

### Datasets

For developing the TzanckNet, we used the cytology archive of Department of Dermatology, Başkent University Medical School, Adana Dr. Turgut Noyan Application and Research Center. This study was approved by Başkent University Institutional Review Board (Project no: KA19/401). There were 2260 Tzanck smear images from erosive-vesiculobullous diseases (pemphigus, herpetic infection, impetigo, Hailey-Hailey disease, contact dermatitis, vesiculobullous dermatophytic infection) collected between December 2006 – December 2019. This dataset is called the *development dataset*. It was used to train and fine tune the model. Afterwards, the finalized model was evaluated using a separate dataset that was collected during January 2020 in the same clinic. It contained 359 images from patients that were not in the development dataset. This is called the *validation dataset*. The images in this dataset were obtained from 15 patients (9 females, 6 males) with ages ranging from 2 to 68 years (34 years on average).

### Dataset preparation

Cytological specimens were stained with May-Grünwald-Giemsa (Bio-Optica), examined microscopically and photographed with a digital camera. Cytological photos taken at ×1000 magnification. Two dermatologists (M.D. and A.H.E.) prepared the dataset with two different microscopes and with two different digital cameras. There were no missing data in the datasets. Images with artifacts (overlapping cells, scratches etc.) were not removed from the dataset. Only images that were blurry at a level that would make them useless in a clinical setting were discarded and the samples were imaged again. Images were labeled, de-identified and given to the data scientist (M.A.N.).

### Reference standard

Images were labeled by two dermatologists (M.D. and A.H.E.). M.D. has 13 years and A.H.E. has 2 years of previous cutaneous cytology experience. Labels were decided by adjudication^32^ i.e. disagreements were resolved through discussion. The image was labeled positively for a cell only if that cell was entirely present in the image.

### Machine learning model development

A deep learning library, fastai^33^, was used to develop the TzanckNet. It is a convolutional neural network (ResNet-50), pretrained on ImageNet. Then, the final classification layer was replaced with six output nodes and retrained. Six output nodes in the final layer correspond to the six cell types we chose our model to recognize: acantholytic cells, eosinophils, hypha, multinucleated giant cells, normal keratinocytes and tadpole cells (Figure 1). An image can contain any combination of these cells; therefore, TzanckNet was designed such that it can output any number/combination of cells for one input image. Each output node, outputs a value between 0 and 1, corresponding to the probability of that cell existing in the input image. Depending on the discrimination threshold this probability is converted to 1 or 0 indicating whether the cell type exists in the image or not. For this study, the discrimination threshold was kept constant as 0.5, except for plotting the receiver operating characteristic curve. The development dataset was used at this stage, for training and tuning the model.

### Statistical analysis

Discrimination and calibration performance were used to assess the TzanckNet. The following metrics were used for discrimination: accuracy, sensitivity, specificity, positive predictive value, negative predictive value and F_1_ score with a discrimination threshold of 0.5. Additionally, receiver operating characteristic curve (ROC curve) was plotted to evaluate the model at different discrimination threshold values. Area under this curve (AUC) was calculated and reported as well. Calibration curve was used to evaluate how well predicted probabilities approximated the actual event probabilities. The validation dataset was used at this stage for evaluating the model. Finally, TzanckNet predictions on selected images were demonstrated.

## Results

Using a convolution neural network, we created a machine learning model called TzanckNet for recognizing six cell types in a Tzanck Smear image. An image can contain multiple cell types, for this reason we designed the model such that it can output more than one cell type. Given one input image, the model outputs six probabilities, corresponding to the predicted probabilities of the existence of six cell types. We fixed a discrimination threshold of 0.5 such that if the predicted probability for any cell type was above this threshold, the model predicted that this cell type existed in the image. These predictions were compared to the reference standard for evaluating the model performance.

The TzanckNet, trained and tuned on the development set, was prospectively validated using 359 images collected in a real-world setting. For each image it made six predictions hence it made 2154 predictions in total. The overall accuracy of the TzanckNet on this dataset was 94.3 % (95% CI 93.4 to 95.3), the sensitivity was 83.7 % (95% CI 80.3 to 87.0) and the specificity was 97.3 % (95% CI 96.5 to 98.1). Results of the other discrimination metrics and performance for each cell are given in Table 1.

**Table 1.**
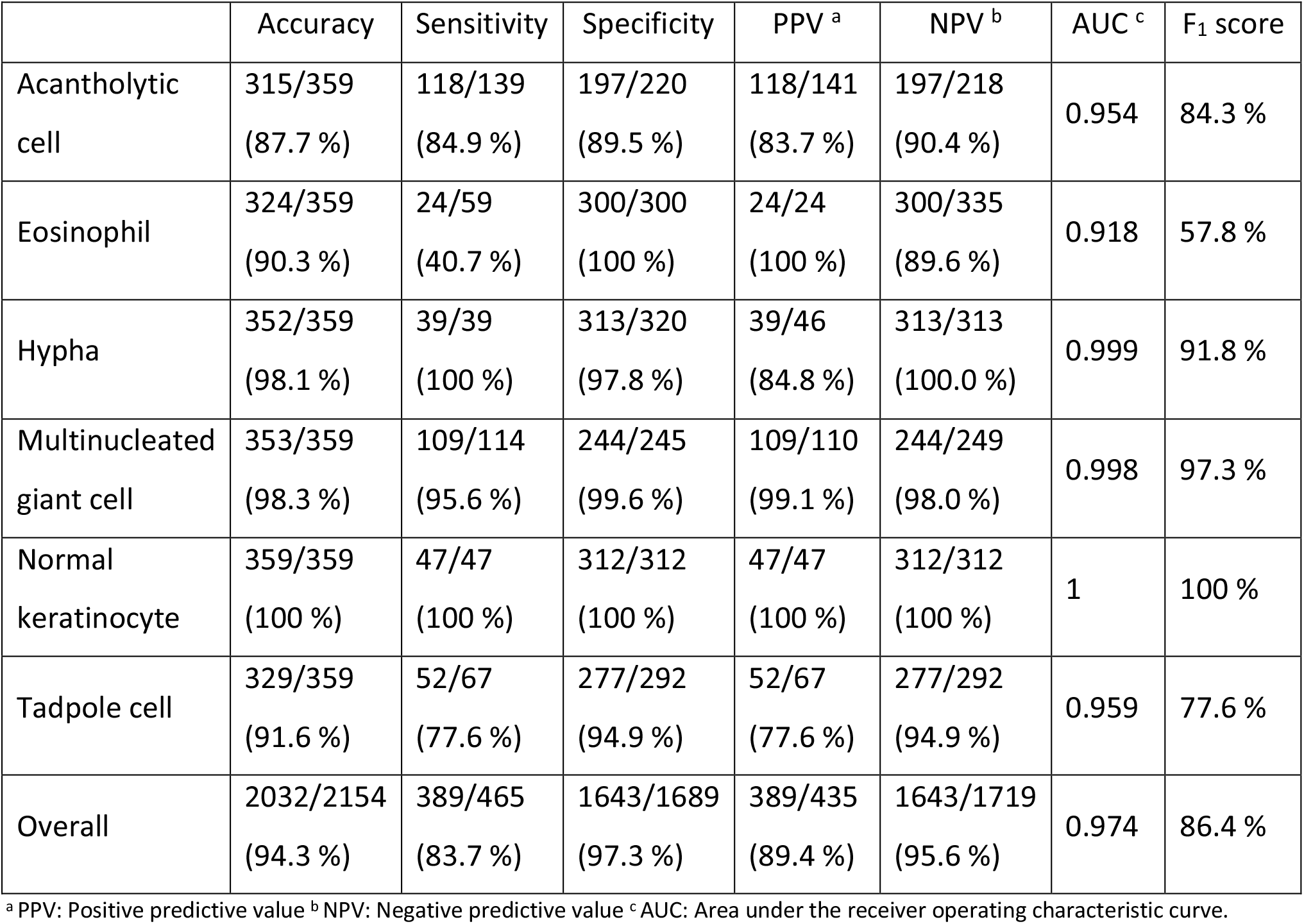
Discrimination metrics of the TzanckNet on the validation dataset that contains 359 Tzanck smear findings.

|  | Accuracy | Sensitivity | Specificity | PPV <sup>a</sup> | NPV <sup>b</sup> | AUC <sup>c</sup> | F<sub>1</sub> score |
| --- | --- | --- | --- | --- | --- | --- | --- |
| Acantholytic cell | 315/359<br>(87.7 %) | 118/139<br>(84.9 %) | 197/220<br>(89.5 %) | 118/141<br>(83.7 %) | 197/218<br>(90.4 %) | 0.954 | 84.3 % |
| Eosinophil | 324/359<br>(90.3 %) | 24/59<br>(40.7 %) | 300/300<br>(100 %) | 24/24<br>(100 %) | 300/335<br>(89.6 %) | 0.918 | 57.8 % |
| Hypha | 352/359<br>(98.1 %) | 39/39<br>(100 %) | 313/320<br>(97.8 %) | 39/46<br>(84.8 %) | 313/313<br>(100.0 %) | 0.999 | 91.8 % |
| Multinucleated giant cell | 353/359<br>(98.3 %) | 109/114<br>(95.6 %) | 244/245<br>(99.6 %) | 109/110<br>(99.1 %) | 244/249<br>(98.0 %) | 0.998 | 97.3 % |
| Normal keratinocyte | 359/359<br>(100 %) | 47/47<br>(100 %) | 312/312<br>(100 %) | 47/47<br>(100 %) | 312/312<br>(100 %) | 1 | 100 % |
| Tadpole cell | 329/359<br>(91.6 %) | 52/67<br>(77.6 %) | 277/292<br>(94.9 %) | 52/67<br>(77.6 %) | 277/292<br>(94.9 %) | 0.959 | 77.6 % |
| Overall | 2032/2154<br>(94.3 %) | 389/465<br>(83.7 %) | 1643/1689<br>(97.3 %) | 389/435<br>(89.4 %) | 1643/1719<br>(95.6 %) | 0.974 | 86.4 % |

<sup>a</sup> PPV: Positive predictive value <sup>b</sup> NPV: Negative predictive value <sup>c</sup> AUC: Area under the receiver operating characteristic curve.

In order to visualize model performance across different discrimination thresholds, overall receiver operating characteristic curve of the network is given in Figure 2a. The area under this curve (AUC) was 0.974. For assessing the performance of the predicted probabilities, calibration curve is given in Figure 2b.

**Figure 2.**
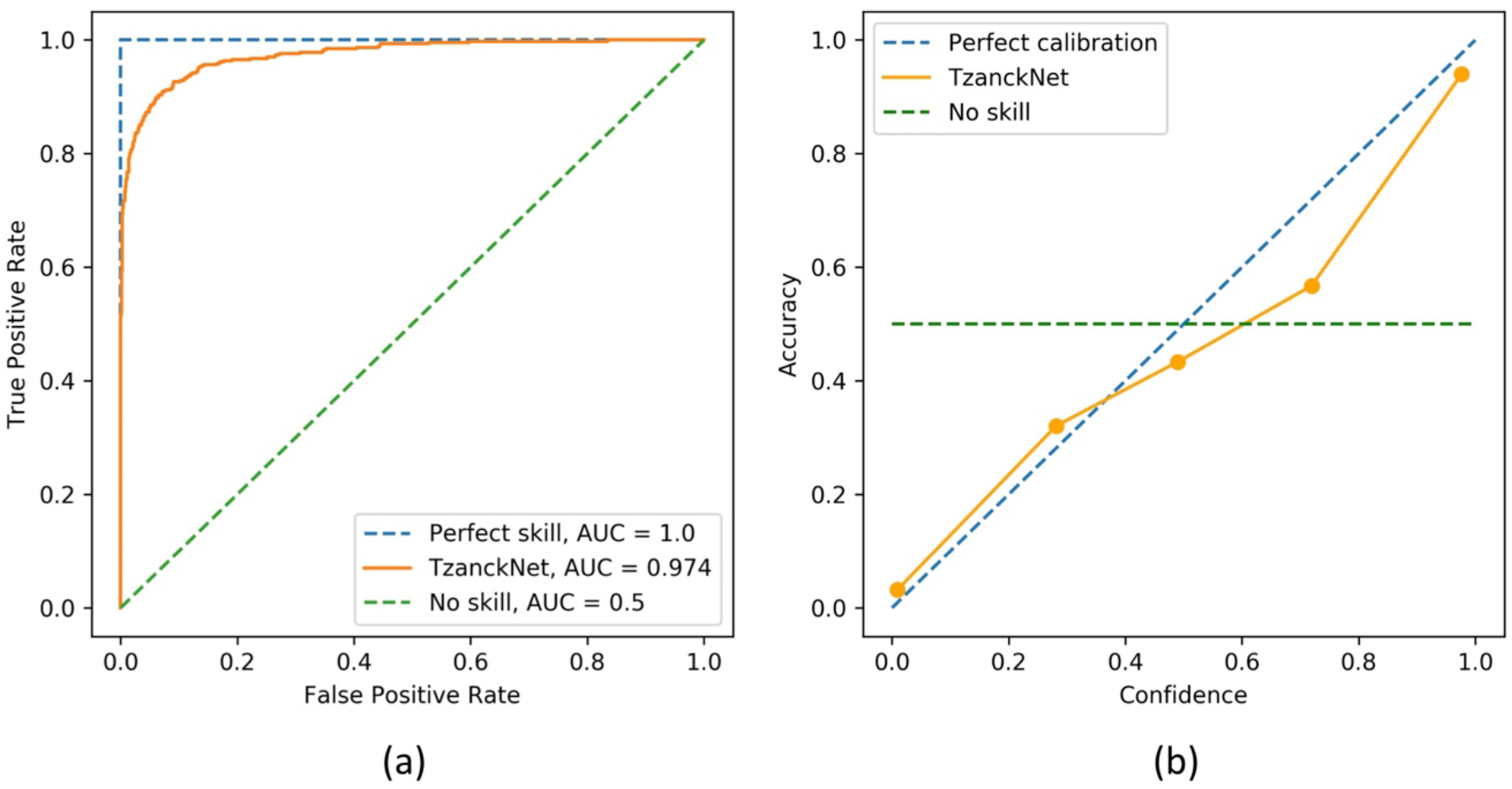
(a) Receiver operating characteristic (ROC) and (b) calibration curves of the TzanckNet on the validation set.

TzanckNet performance on selected Tzanck smear findings are given in Figure 3. Figure 3a contains a multinucleated giant cell (arrow). At first sight during labeling, it seemed like the cell had one circular nucleus and was hence labeled as an acantholytic cell. Upon further examination, it was realized that the cell actually had three nuclei instead of one and the label was changed to a multinucleated giant cell. The TzanckNet recognized that there was as a multinucleated giant cell, but it also gave a 44.6 % probability for the existence of an acantholytic cell. Two eosinophils can be seen in Figure 3b (arrows), however, TzanckNet didn’t recognize them. In Figure 3c, only some part of an acantholytic cell (arrow) is visible inside the image therefore the image was labelled as having no acantholytic cell in the image. TzanckNet predicted that there was an acantholytic cell, but it was not confident (51.1 % predicted probability). Figure 3d contains an atypical acantholytic cell (arrow). It is atypical because the perinuclear halo is not evident and in some parts cytoplasm is not discernible. Therefore, the image was labeled as not having an acantholytic cell. The TzanckNet, however, detected that there is an acantholytic cell in this image, and it was 86.3 % confident.

**Figure 3.**
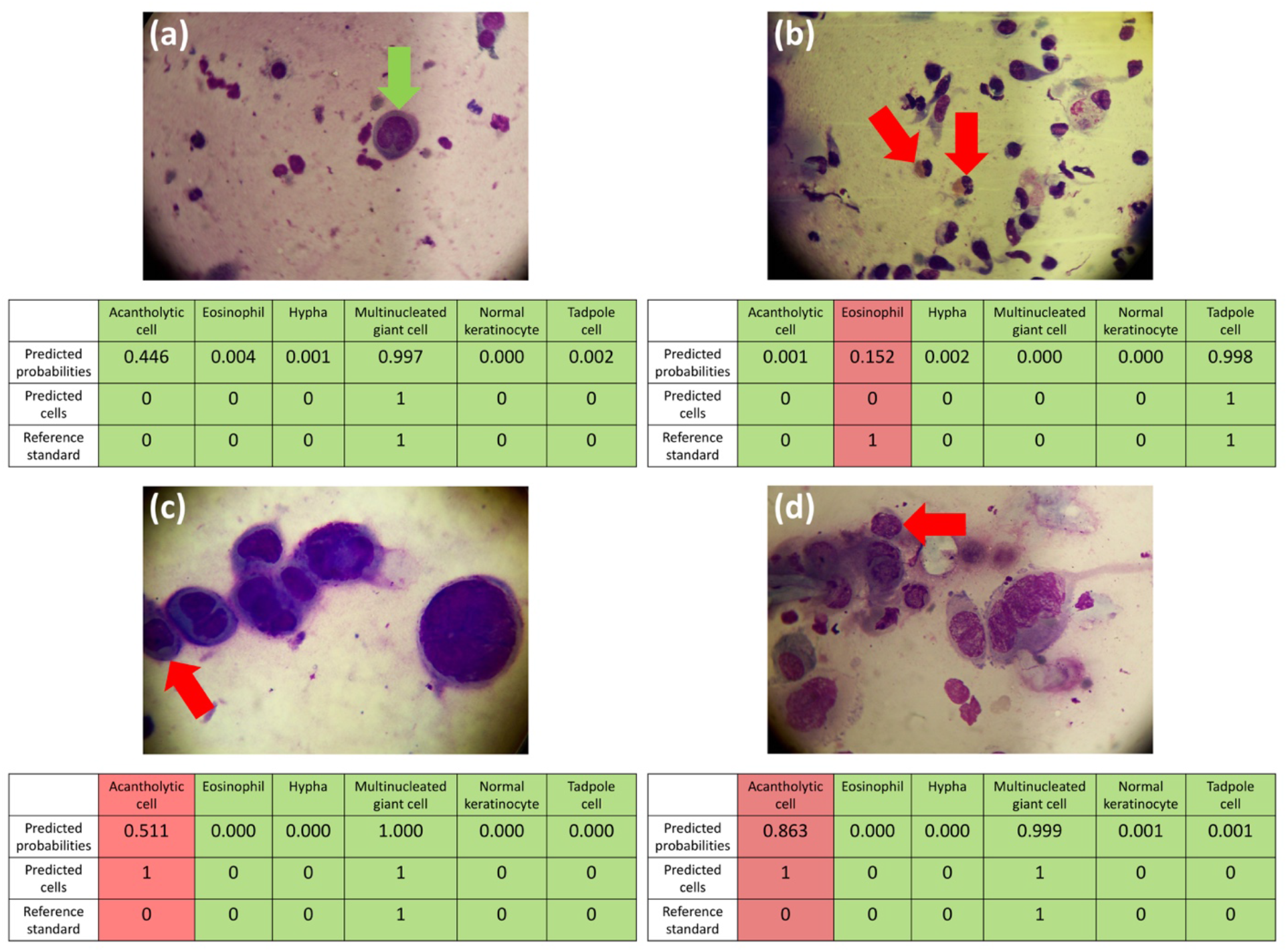
TzanckNet predictions and the corresponding reference standards for four selected images. For each cell type and image, TzanckNet predicts the probability of that cell type being present in the image. Probabilities are then converted to decisions of absence (0) or presence (1) of that cell, using a discrimination threshold of 0.5. Decisions matching with the reference standards are marked with green, and with red otherwise. The red arrows indicate the cells that are related to the false predictions. The green arrow indicates a multinucleated giant cell.

## Discussion

Tzanck smear test is an inexpensive test that can provide rapid diagnosis of many dermatological diseases. Interest in Tzanck smear test has increased in recent years. Nevertheless, in most clinics, this simple test is either not performed or is only used to diagnose a few diseases, mainly due to lack of experience. In a dermatology clinic where 75 patients for whom no diagnosis could be established via face-to-face clinical examinations and cytological evaluations, telemedicine was used to overcome this issue.^34^ Although this is a reasonable solution, the necessary expertise is still scarce, scheduling is necessary, and one can only examine a limited number of images during a session. In this work we demonstrated that TzanckNet can identify six cell types in Tzanck smear findings. It can analyze hundreds of images in a minute with high accuracy.

The overall performance of the TzanckNet shows that deep learning has the potential to aid dermatologists for analyzing Tzanck smear findings. It was able to recognize cells with high accuracy. Moreover, the calibration performance shows that the predicted probabilities can also help with the interpretation of images. For example, if a cell type exists with 99 % estimated probability the network is almost certain that this cell is present in the image whereas if the estimated probability is 70 % the network tends to predict that this cell is present but not confidently. If the network is not confident about its predictions, additional images can be taken.

Examining individual predictions can provide a better understanding of the model’s strengths and limitations hence indicating ways to improve it furthermore. For example, TzanckNet was able to recognize correctly a multinucleated cell that looks similar to an acantholytic cell (Figure 3a). Looking at Table 1, low sensitivity on eosinophils draws attention. Specificity being 100 % indicates that all of the errors for eosinophils were false negatives, there were no false positives. Individual examination of the images that resulted in false negatives revealed that they had less than five eosinophils on average whereas the examination of true positives revealed that they had more than ten eosinophils on average. One of the images that resulted in a false negative eosinophil can be seen in Figure 3b. Indeed, it contains only two eosinophils (arrows). This suggests that for TzanckNet to recognize eosinophils, they should be abundant in the image. Figure 3c and 3d indicates model performance may not be ideal for cells that are partially in the image or atypical cells.

The most common use of Tzanck smear test is the diagnosis of herpetic infection.^7,35^ Herpetic infections may be primary or develop on other dermatoses. Early diagnosis of herpetic infections is very important in terms of early treatment and prevention of the spread of the disease. When Tzanck smear is not performed, patients receive unnecessary treatment or more complex diagnostic methods such as polymerase chain reaction or histopathological examinations are requested.^2^ Tzanck smear test is cheaper and faster than both of these methods.^4^ The characteristic finding of herpetic infection in the Tzanck smear test is acantholytic cells with multinuclear giant cells.^2^ Another disease that is related to these cells is pemphigus. It is a rare and severe autoimmune disease that can cause mortality. Tzanck smear test is not only important in early diagnosis of the disease but also in the diagnosis of recurrent lesions. A definitive diagnosis of pemphigus requires histopathological examination and immunofluorescence test. However, it is not possible to take a biopsy each time for recurrent lesions. Herpetic infection and candidiasis should be excluded ruled out especially in newly developed lesions resistant to corticosteroid treatment in oral mucosa. If this test is not performed, patients will receive unnecessary steroid treatment, which may lead to sepsis.^36^ In pemphigus patients, unlike herpetic infections, acantholytic cells without multinuclear giant cells are observed in cytological examination.^2,9^ In this study, TzanckNet showed that it was able to recognize acantholytic and multinuclear giant cells demonstrating its diagnostic potential for suspected herpetic infection and pemphigus.

A few tadpole cells are observed in all vesiculobullous diseases. However, numerous tadpole cells are characteristic cytologic findings of spongiotic dermatitis^37^ and eczema. Tzanck smear is used for two purposes in eczema patients. The first is to distinguish eczema from other acantholytic diseases and the second is to detect bacterial, fungal and viral infections. Tzanck smear test performed for both purposes prevent unnecessary treatment. If the test is not done, infections can spread easily with steroid treatments used for eczema. Accuracy of the model on tadpole cells was 91.6 %, however it should be noted that the specificity was high, but sensitivity was low suggesting that the model tends to give false negatives.

Cutaneous eosinophilic infiltration may develop due to various causes such as infectious, inflammatory and neoplastic diseases. Histopathology is often used in the differentiation of these diseases. However, cytological examination has been used in only a few studies focused on the utility of cytology in some eosinophilic diseases. Besides detecting eosinophils, cytology can also show the various infectious etiologic agents and distinguish some inflammatory diseases. If eosinophils are present, bacterial and fungal structures should be investigated. The performance of TzanckNet on eosinophils and hypha suggests that the network can also be utilized as a decision support tool for diagnosing aforementioned diseases.

The overall performance of the TzanckNet demonstrates its strong diagnostic potential. We would like to emphasize that we propose this model as a clinical decision support system to improve physician accuracy and efficiency in the clinical workflow, not as a replacement or a tool that can directly output a diagnosis. It has been shown that human-machine combination works better than either alone.^38–40^ Additionally, TzanckNet can be used to train dermatologists on Tzanck smear test.

### Limitations

Like all methods/models, explicitly reporting known limitations of TzanckNet is crucial for clinicians and researchers who want to use and improve it. The model was trained using Tzanck smear findings of patients from Turkey, stained with May-Grünwald-Giemsa and taken with ×1000 magnification. The performance of the model on other populations, sample preparation procedures and imaging systems remains unknown at this point. Model performance on cells that are atypical or that overlap or appear partially in the image may not be ideal. Finally, yet importantly, the model can recognize which cell types exist in an image, but it cannot localize the cells. Localizing the cells has many advantages such as being able to count the cells and being able to say precisely which cells in the image are recognized and what their types are. Two major improvements are needed to realize the full potential of the TzanckNet: the first is to retrain and validate the model with other populations, stains, and magnifications for better generalization, and the second is to add object detection to the model for localizing the cells.

Nevertheless, this study showed that TzanckNet can analyze the cytological findings of erosive-vesiculobullous diseases accurately. The model can be extended for analyzing granulomatous and tumoral diseases.

## Conclusions

Tzanck smear test is a valuable but underappreciated diagnostic tool. This work introduced TzanckNet, a machine learning model that can analyze Tzanck smear findings with high accuracy. It can be used as a clinical decision support system as well as a training tool for new physicians. It has the potential to spread the use of Tzanck smear test, decrease the number of biopsies, prevent unnecessarily long antibiotic treatments, help early diagnoses for fatal diseases, decrease costs, and thus improve patient well-being.

## Author contributions

M.A.N. and M.D. had full access to all of the data in the study and take responsibility for the integrity of the data and the accuracy of the data analysis. M.A.N. and M.D. proposed the study and its design. M.D. and A.H.E. prepared, imaged, and labeled the Tzanck smears. M.A.N. developed the machine learning model and wrote the manuscript. All authors reviewed, revised, and approved the final manuscript.

## Funding

None

## Conflict of Interest Disclosures

None

## Data Availability

The datasets generated and analyzed during the study are not currently publicly available.

## Notes

### Competing Interest Statement

The authors have declared no competing interest.

### Funding Statement

No external funding.

### Author Declarations

This study was approved by Başkent University Institutional Review Board (Project no: KA19/401).

